# COVID-19 serial interval estimates based on confirmed cases in public reports from 86 Chinese cities

**DOI:** 10.1101/2020.04.23.20075796

**Authors:** Zhanwei Du, Xiaoke Xu, Ye Wu, Lin Wang, Benjamin J. Cowling, Lauren Ancel Meyers

## Abstract

As a novel coronavirus (COVID-19) continues to spread widely and claim lives worldwide, its transmission characteristics remain uncertain. Here, we present and analyze the serial intervals–the time period between the onset of symptoms in an index (infector) case and the onset of symptoms in a secondary (infectee) case–of 339 confirmed cases of COVID-19 identified from 264 cities in mainland China prior to February 19, 2020. Here, we provide the complete dataset in both English and Chinese to support further COVID-19 research and modeling efforts.

## Background & Summary

Key aspects of 2019 novel Coronavirus Disease (COVID-19) transmission mechanisms remain unclear (1). By April 22, 2020, there have been 2,471,136 confirmed cases and 169,006 deaths in 210 countries (2), while key aspects of the transmission dynamics of COVID-19 remain unclear (3). The COVID-19 serial interval is defined as the period of time between a primary case-patient (infector) with symptom onset and a secondary case-patient (infectee) with symptom onset (4,5). Obtaining reliable estimates for the distribution of COVID-19 serial intervals is a crucial input in deciding the specific number of reproductions (R_0_), which can demonstrate the magnitude of the measures needed to contain an epidemic (6). This quantity can not however be deduced from the regular case count data alone (7). Recent estimates for the mean serial interval of COVID-19 range from 7.5 days [95% CI 5.3-19] (13) to 4.0 days [95% CrI 3.1-4.9] (17) based on data from 6 and 28, respectively.

We have curated a detailed dataset of 339 COVID-19 translated from case reports posted online by 18 provincial health departments in mainland China outside of Hubei Province between January 20 and February 19, 2020 (Appendix Table 1). Each report consists of a probable date of initiation of symptoms for both the infector and the infectee, as well as the probable locations of infection for both case-patients. Based on these reports, we estimate that COVID-19 has an average serial interval of 5.29 days (95% CI 4.72–5.86 days).

## Methods

### Data

We obtained publicly available data from 264 cities in mainland China on 9,120 reported COVID-19 infection events, which were accessible online as of February 19, 2020. The data were collected from the websites of the provincial departments of public health and translated into English from Chinese (Appendix Table 1).We then searched the data for clearly identified transmission events that consisted of: (i) a known infector and infectee, (ii) recorded infection locations for both cases, and (iii) documented symptom onset dates and locations for both cases. We thereby obtained 339 infector-infectee pairs identified via contact tracing in 86 Chinese cities between January 20, 2020 and February 19, 2020 (Appendix Figure 1). The index cases (infectors) for each pair are reported as either importations from the city of Wuhan (N = 136), importations from cities other than Wuhan (N = 45) or local infections (N = 158). The cases included 556 distinct individuals, with 62 index cases infecting multiple individuals and 18 individuals occurring as both infector and infected individuals. We range from 0 to 86 years of age, which include 250 women and 306 males.

### Estimating Serial Interval Distribution

For each infector-infectee pari, we measured the number of days between the reported symptom onset date of the infector and the recorded symptom onset date of the infectee. Negative values suggest that the infectee developed symptoms before the infector. We then used the fitdist function in Matlab (8) to fit a normal distribution to all 339 observations. It provides reliable estimates of the mean and standard deviation, with 95% confidence intervals. We applied the same method to estimate the means and standard deviations after stratifying by whether the index case was infected locally or imported.

### Model comparison

We used maximum likelihood fitting and the Akaike information criterion (AIC) to evaluate four candidate models for the COVID-19 serial interval distributions: normal, lognormal,

Weibull and gamma. Since our serial interval data includes a substantial number of non-positive values, we fit the four distributions both to truncated data in which all non-positive values are removed and to shifted data in which nine days are added to each observation (Appendix Figure 1, Appendix Tables 1,3). The lognormal distribution provides the best fit for the truncated data (followed closely by the gamma and Weibull). However, we do not believe there is cause for excluding the non-positive data and would caution against making assessments and projections based on the truncated data. The normal distribution provides the best fit for the full dataset (shifted or not) and thus is the distribution we recommend for future epidemiologic assessments and planning.

### Distribution analysis

To facilitate interpretation and future analyses, we summarize key characteristics of the COVID-2019 infection report dataset.

#### Age Distribution

Of the 556 unique cases in the dataset, 1.3%, 2.2%, 51.08%, 32.01% and 13.49% were ages 0–4, 5–17, 18–49, 50–64, and over 65 years, respectively. Across all transmission events, approximately one third occurred between adults ages 18 to 49, ∼94% had an adult infector (over 18), and 100% had an adult infectee (over 18) (Appendix Table 4).

#### Secondary Case Distribution

Across the 339 transmission events, there were 240 unique infectors. The mean number of transmission events per infector is 1.41 (Appendix Figure 2) with a maximum of 7 secondary infections reported from a 80 year old male in Cangzhou city of Hebei Province.

#### Geographic Distribution

The 339 transmission events were reported from 86 Chinese cities in 17 Chinese provinces and Tianjin (Appendix Figure 3). There are 18 cities with at least five infection events and 68 cities with fewer than five infection events in the sample. The maximum number of reports from a city is 36 for Xinyang, which reported 269 cumulative cases as of February 19, 2020.

## Data Records

This dataset is released by one comma-separated values (CSV) file, which has been uploaded to github (MeyersLabUTexas/COVID-19). In the csv file, there are 21 columns ordered by index infection, secondary infection and source. The format for this file is the following.

- Event index: unique identifier for each infection event.
- Index ID: unique identifier for index reported case.
- Secondary ID: unique identifier for secondary reported case.
- City (Chinese): initial entry of name of the city in Chinese, in which the index and secondary cases are reported.
- City (English): initial entry of name of the city in English, in which the index and secondary cases are reported.
- Index - infection location (Chinese): initial entry of name of the city in Chinese, in which the index case is infected.
- Index-infection location (English): initial entry of name of the city in English, in which the index case is infected
- Index - symptom onset date: date following ISO 8601 format (YYYY-MM-DD) when the index case had symptom onset.
- Index - Age: age of the index case reported in years
- Index - Sex: numerical values for sex (Male as 1 and Female as 2)
- Secondary - infection location (Chinese): initial entry of name of the city in Chinese, in which the secondary case is infected.
- Secondary-infection location (English): initial entry of name of the city in English, in which the secondary case is infected.
- Secondary - symptom onset date: date following ISO 8601 format (YYYY-MM-DD) when the index case had symptom onset.
- Secondary - Age: age of the Secondary case reported in years
- Seconday-Sex: numerical values for sex (Male as 1 and Female as 2)
- Contact type: character values to denote whether the infection is household or not
- URL: URL link of urban Municipal Health Commission for each event
- Data source: name of urban Municipal Health Commission for each event
- Index description (Chinese): description of index case in Chinese from data source.
- Secondary description (Chinese): description of secondary case in Chinese from data source.
- Other description (Chinese): description of other information in Chinese from data source.

## Technical Validation

Thirty-three of the 339 cases suggest that the infectee had earlier symptoms than the infection. Therefore, there may be presymptomatic transmission. Because of these negative serial intervals, the COVID-19 serial intervals are better fit by normal distributions than the generally assumed gamma or Weibull distributions (10,11), which are limited to positive values (Appendix). Assuming a normal distribution, we predict a mean serial interval for COVID-19 of 5.29 days (95% CI 4.72–5.86) with an SD of 5.32 days (95% CI 4.95–5.75) (**Figure 1**), which is significantly lower than the mean serial intervals of 8.4 days recorded for extreme acute respiratory syndrome (11) and 12.6 days (12) to 14.6 days (13) for Middle East respiratory syndrome. The mean serial interval is slightly longer when the index case is imported (5.61 days [95% CI 4.83–6.40]) versus locally infected (4.92 days [95% CI 4.09–5.75]), but marginally shorter when the secondary transmission occurs inside the household (4.57 days [95% CI 3.76–5.38]) versus outside the household (5.85 days [95% CI 5.06–6.64]).

**Figure 1.**
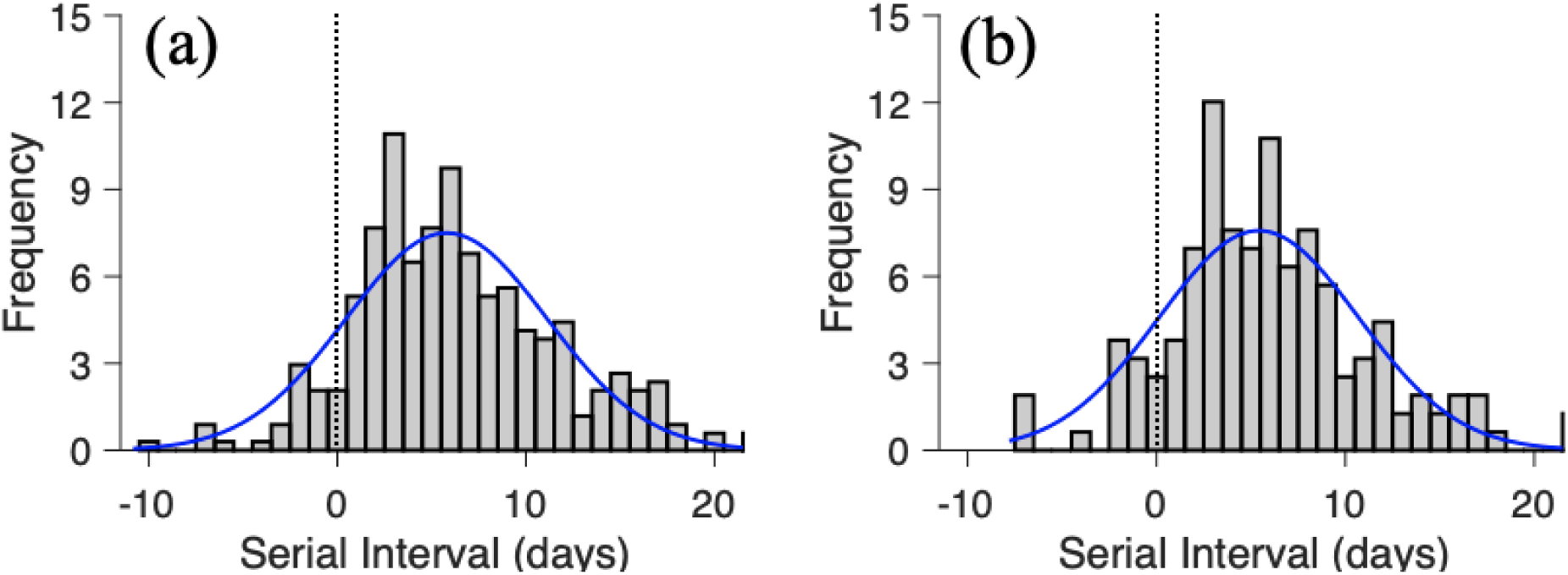
Estimated serial interval distribution for 2019 novel coronavirus disease (COVID-19) based on 339 reported transmission events in mainland China outside of Hubei Province from January 20 to February 19, 2020. Bars indicate the number of infection events with specified serial interval, and blue lines indicate fitted normal distributions for (A) all infection events (N = 339) reported across 86cities of mainland China as of February 19, 2020, and (B) the subset infection events (n = 158) in which both the infector and infectee were infected in the reporting city (i.e., the index patient’s case was not an importation from another city). Negative serial intervals (left of the vertical dotted lines) suggest the possibility of COVID-19 transmission from asymptomatic or mildly symptomatic case-patients.

These estimates reflect the recorded dates of onset of symptoms for 556 case-patients from 86 China cities, ranging from 0 to 86 years of age (mean 46.5 years, SD 16.2 years). Other recent estimates for the average COVID-19 serial interval based on case data from mainland China, Taiwan, Hong Kong, Vietnam, South Korea, Germany, and Singapore include 7.5 days (95% CI 5.3–19) [13], 4.4 days (95% CI 2.9–6.7) (17), and 4.0 days [95% CrI 3.1–4.9] (18), based on considerably smaller samples of 6, 21, and 28 infector–infectee pairs, respectively. Although none of these studies indicate negative serial intervals before the infector in which the infectee had symptoms, 9.73% of the serial intervals in our study are negative.

We note in our estimates four possible causes of bias (19). First, the data is restricted to online records of reported cases and thus could be skewed towards more serious cases in areas with high-functioning healthcare and public health systems. The rapid isolation of these case-patients may have avoided longer serial intervals, possibly moving our estimate downward relative to the serial intervals that could be found in an uncontrolled outbreak.

Second, the distribution of serial intervals varies during an outbreak, with time contracting around the outbreak peak between successive cases (20). A susceptible person would possibly get infected faster if they are surrounded by two rather than one infected person. Since our projections are mainly based on transmission events recorded during the early stages of outbreaks, we do not specifically account for such fragmentation and view the figures at the start of an epidemic as simple serial intervals. However, if any of the recorded infections occurred in the midst of increasing clusters of cases, our estimates may represent successful (compressed) serial intervals anticipated during an epidemic growth period. Third, each infector’s identity and timing of the onset of symptoms is probably based on an individual memory of past events. If the precision of the recall is impeded by time or trauma, case-patients may be more likely to relate infection over prior experiences (longer serial intervals) to recent experiences (short serial interval). By comparison, the recorded serial intervals may be skewed upwards by travel-related transmission delays from primary case patients infected in Wuhan or another city before returning home. If their infectious cycle begins while traveling, then we may not be able to detect events of early transmission with shorter serial intervals.

Given the heterogeneity in the nature and reliability of these reports, we caution that our findings should be viewed as working hypotheses concerning COVID-19 infectiousness, which require further testing as more data become accessible.

## Data Availability

Not applicable

## Acknowledgments

We acknowledge the financial support from NIH (U01 GM087719) and the National Natural Science Foundation of China (61773091).

## Appendix

**Appendix Table 1.** Model comparison for COVID-19 serial intervals based on 158 reported transmission events in China between January 20, 2020 and February 19, 2020 in which both the infector and infectee were infected in the reporting city (i.e., the index case was not an importation from another city)

| Data | Distribution | Mean/Shape (95% CI) | SD/Scale (95% CI) | AIC |
| --- | --- | --- | --- | --- |
| Original data | Normal (Mean, SD) | 4.92 (4.09-5.75) | 5.26 (4.74– 5.92) | 976.28 |
| Truncated (>0) | Normal (Mean, SD) | 6.3 (5.54-7.06) | 4.44 (3.96-5.04) | 776.74 |
|  | Lognormal (Shape, Scale) | 1.57 (1.44-1.71) | 0.78 (0.69-0.88) | 732.27 |
|  | Gamma (Shape, Scale) | 2.03 (1.62-2.54) | 3.1 (2.41-4) | 727.6 |
|  | Weibull (Shape, Scale) | 7 (6.21-7.9) | 1.49 (1.31-1.7) | 729.94 |
| Shifted (+9d) | Normal (Mean, SD) | 13.92 (13.09-14.75) | 5.26 (4.74-5.92) | 976.28 |
|  | Lognormal (Shape, Scale) | 2.54 (2.46-2.62) | 0.5 (0.45-0.57) | 1037.33 |
|  | Gamma(Shape, Scale) | 5.49 (4.43-6.8) | 2.54 (2.03-3.17) | 995.39 |
|  | Weibull(Shape, Scale) | 15.56 (14.66-16.51) | 2.77 (2.46-3.12) | 980.2 |

**Appendix Table 2.** Estimated serial interval distributions based on the location of index infection (imported versus local) and the secondary infection (household versus non-household)*

| Group | Mean (95 CI%) | SD (95 CI%) | Proportion of serial intervals <0 |
| --- | --- | --- | --- |
| All (N = 339) | 5.29 (4.72– 5.86) | 5.32 (4.95–5.75) | 9.73% (N = 33) |
| Locally infected index case (n = 158) | 4.92 (4.09-5.75) | 5.26 (4.74–5.92) | 12.03% (N = 19) |
| Imported index case (n = 181) | 5.61 (4.83–6.40) | 5.36 (4.86–5.98) | 7.73% (N = 14) |
| Household secondary infection (n = 149) | 4.57 (3.76–5.38) | 4.98 (4.48–5.62) | 10.07% (N = 15) |
| Non-household secondary infection (n = 190) | 5.85 (5.06–6.64) | 5.52 (5.01–6.14) | 9.47% (N = 18) |

**Appendix Table 3.** Model comparison for COVID-19 serial intervals based on all 339 reported transmission events in China between January 20, 2020 and February 19, 2020.

| Data | Distribution | Mean/Shape (95% CI) | SD/Scale (95% CI) | AIC |
| --- | --- | --- | --- | --- |
| Original data | Normal (Mean, SD) | 5.29 (4.72– 5.86) | 5.32 (4.95– 5.75) | 2,098.35 |
| Truncated (>0) | Normal (Mean, SD) | 6.61 (6.08-7.13) | 4.54 (4.2-4.94) | 1691.81 |
|  | Lognormal (Shape, Scale) | 1.61 (1.52-1.71) | 0.8 (0.74-0.87) | 1621.07 |
|  | Gamma (Shape, Scale) | 1.97 (1.69-2.29) | 3.35 (2.82-3.98) | 1603.23 |
|  | Weibull (Shape, Scale) | 7.34 (6.77-7.96) | 1.5 (1.37-1.64) | 1603.89 |
| Shifted (+9d) | Normal (Mean, SD) | 17.29 (16.72-17.86) | 5.32 (4.95-5.75) | 2098.35 |
|  | Lognormal (Shape, Scale) | 2.8 (2.76-2.83) | 0.36 (0.34-0.39) | 2168.09 |
|  | Gamma(Shape, Scale) | 9.29 (8.01-10.77) | 1.86 (1.6-2.17) | 2117.75 |
|  | Weibull(Shape, Scale) | 19.18 (18.57-19.81) | 3.47 (3.2-3.76) | 2106.68 |

**Appendix Table 4.** Age distribution for the 339 infector–infectee pairs. Each value denotes the number of infector-infectee pairs in the specified age combination. Age was not reported for the remaining 11 pairs.

| Infector | Infectee |  |  |  |  | Total |
| --- | --- | --- | --- | --- | --- | --- |
|  | 0–4 | 5–17 | 18–49 | 50–46 | ≥65 |  |
| 0–4 | 0 | 0 | 0 | 0 | 0 | 0 |
| 5–17 | 0 | 0 | 0 | 0 | 0 | 0 |
| 18–49 | 6 | 10 | 100 | 51 | 27 | 194 |
| 50–46 | 1 | 3 | 44 | 44 | 12 | 104 |
| ≥65 | 0 | 0 | 19 | 11 | 11 | 41 |
| Total | 7 | 13 | 163 | 106 | 50 | 339 |

**Appendix Table 5.** Case report data for 339 COVID-19 infections occurring in 86 Chinese cities by February 19, 2020. Data iswill be available upon publication at https://github.com/MeyersLabUTexas/COVID-19.

**Appendix Figure 1.**
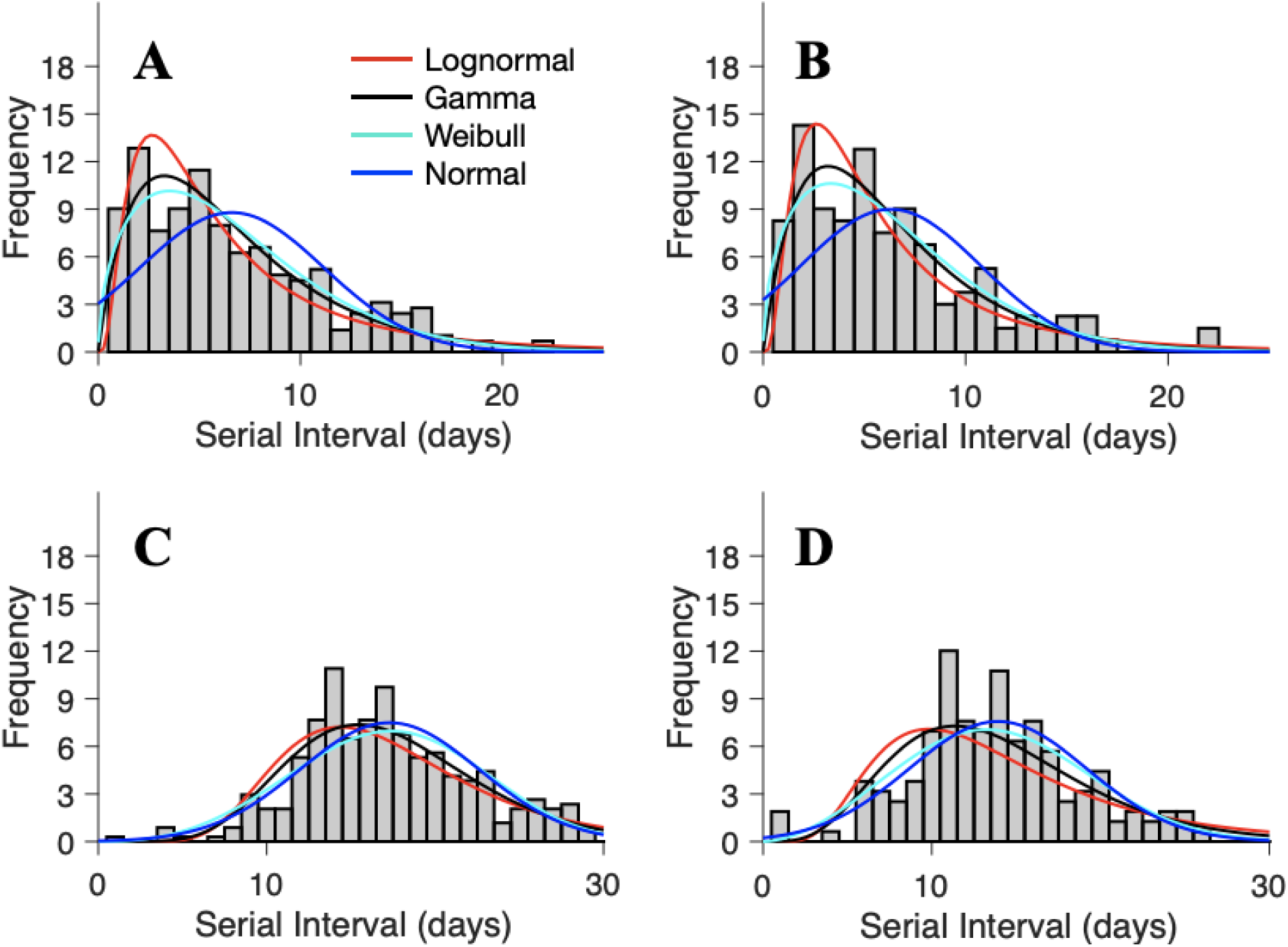
Maximum likelihood distributions fit to transformed COVID-19 serial intervals (339 reported transmission events across 86 cities in Mainland China between January 20, 2020 and February 19, 2020). To evaluate several positive-valued distributions (lognormal, gamma and Weibull), we took two approaches to addressing the negative-valued data. First, we left truncated the data (i.e., removed all non-positive values) for (a) all 339 infection events and (b) the subset of infection events (N = 158) in which both the infector and infectee were infected in the reporting city (i.e., the index case was not an importation from another city). Second, we shifted the data by adding nine days to each reported serial interval for (c) all infection events and (d) the subset of infection events in which both the infector and infectee were locally infected. Bars indicate the number of infection events with the specified serial interval and colored lines indicate the fitted distributions. Parameter estimates and AIC values are provided in Table S4.

**Appendix Figure 2.**
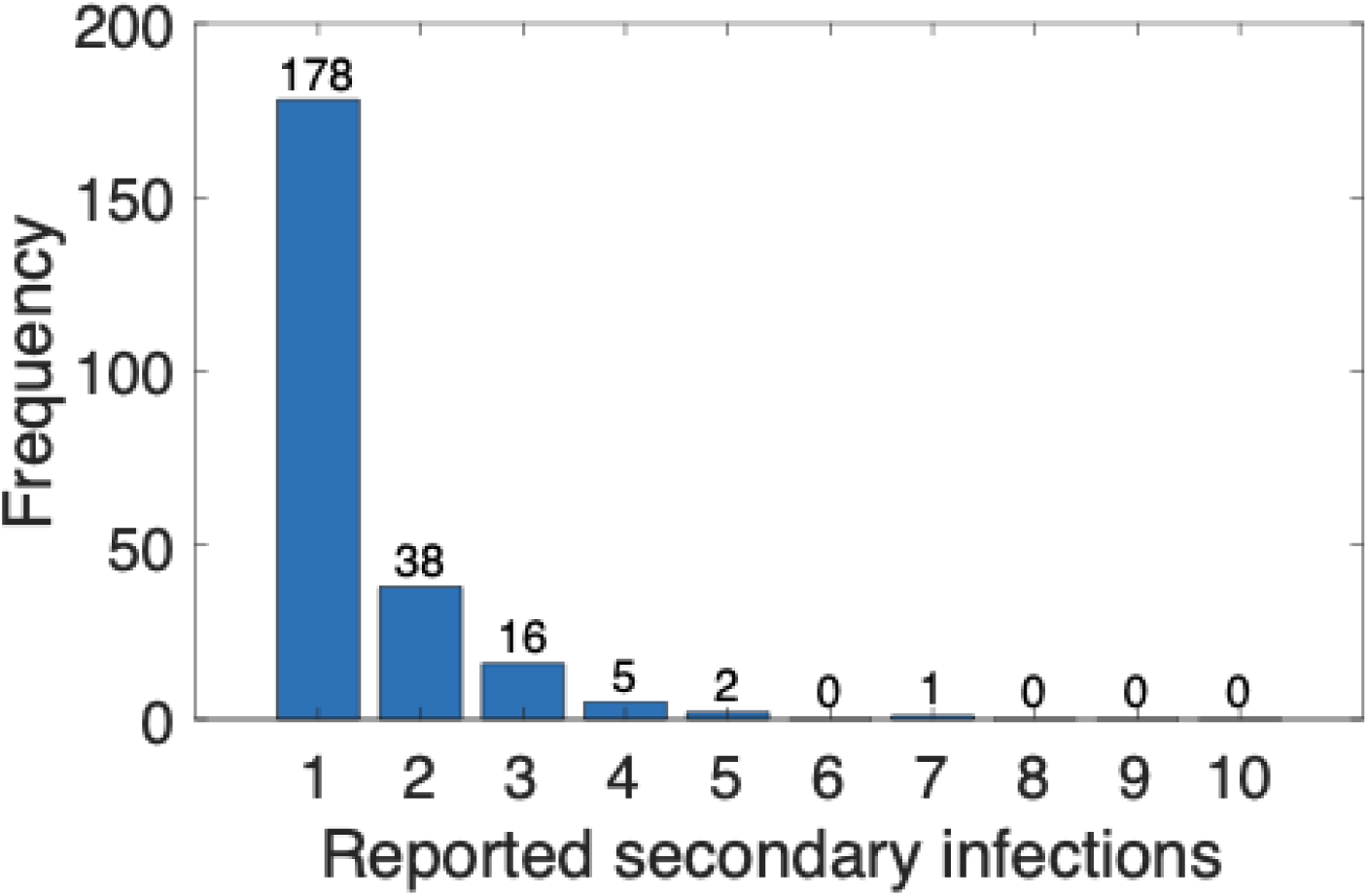
Number of infections per unique index case in the infection report dataset. There are 240 unique infectors across the 339 infector-infectee pairs. The number of transmission events reported per infector ranges from 1 to 7, with ∼74% having only one.

**Appendix Figure 3.**
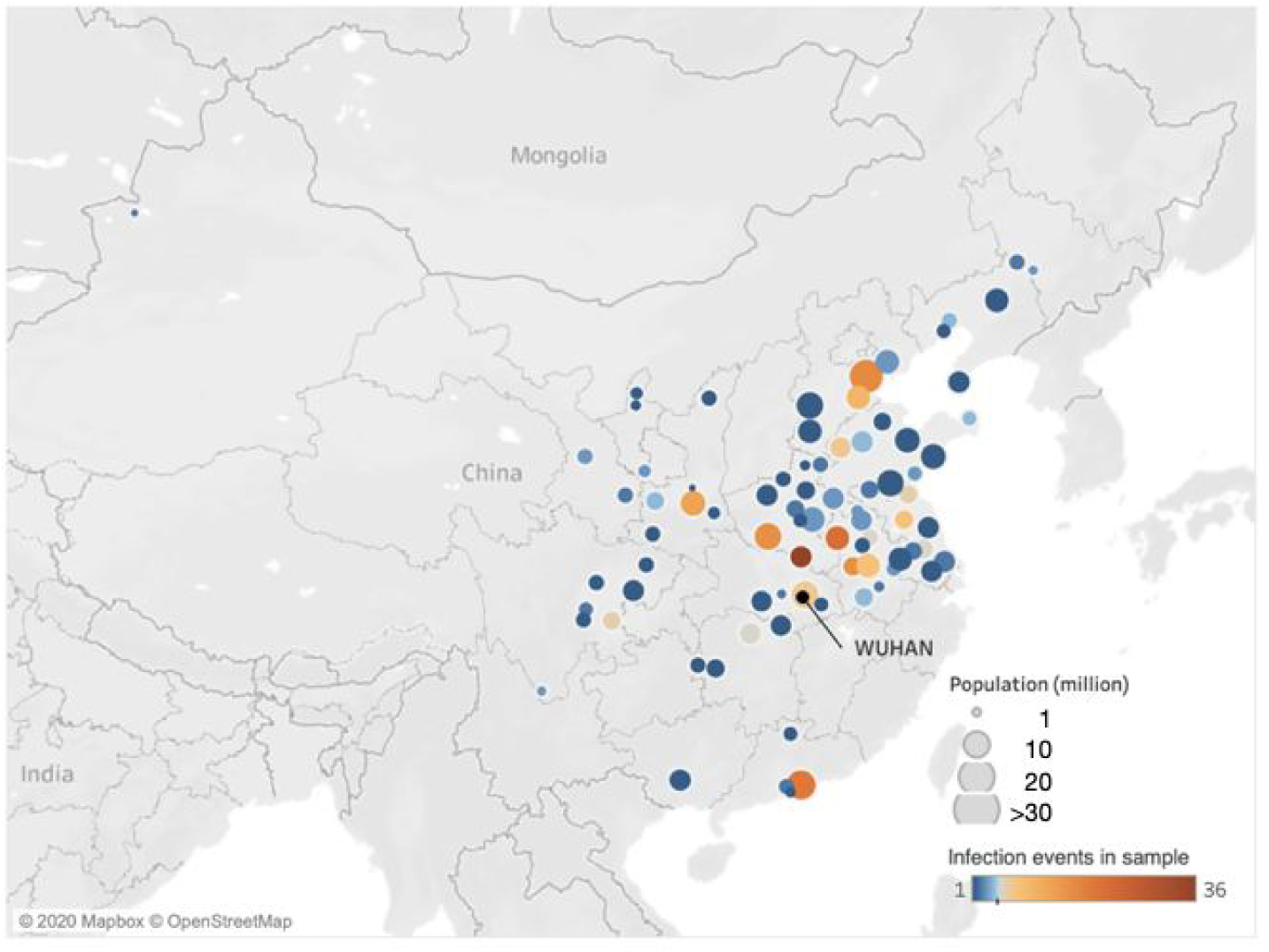
Geographic composition of the infection report dataset. The data consist of 339 infector-infectee pairs reported by February 19, 2020 across 86 cities in mainland China. Colors represent the number of reported events per city, which range from 1 to 36, with an average of 3.94 (SD 5.65) infection events. The 68 cities with fewer than five events are colored in blues; the 18 cities with at least five events are colored in shades of orange.

## Code Availability

Matlab code for data analysis of location correction and mobility network construction can be obtained freely by contacting the first author with no restrictions to access.

## References

1. WHO | Pneumonia of unknown cause – China. 2020 Jan 30 [cited 2020 Feb 18]; Available from: https://www.who.int/csr/don/05-january-2020-pneumonia-of-unkown-cause-china/en/

2. World Health Organization. Coronavirus disease 2019 (COVID-19): situation report, 93 [Internet]. Available from: https://www.who.int/docs/default-source/coronaviruse/situation-reports/20200422-sitrep-93-covid-19.pdf?sfvrsn=35cf80d7_4

3. Cowling BJ, Leung GM. Epidemiological research priorities for public health control of the ongoing global novel coronavirus (2019-nCoV) outbreak. Euro Surveill [Internet]. 2020 Feb 13; Available from:http://dx.doi.org/10.2807/1560-7917.ES.2020.25.6.2000110

4. Giesecke J. Modern infectious disease epidemiology. CRC Press; 2017.

5. Svensson A. A note on generation times in epidemic models. Math Biosci. 2007 Jul;208(1):300–11.

6. Wallinga J, Lipsitch M. How generation intervals shape the relationship between growth rates and reproductive numbers. Proc Biol Sci. 2007 Feb 22;274(1609):599–604.

7. Vink MA, Bootsma MCJ, Wallinga J. Serial Intervals of Respiratory Infectious Diseases: A Systematic Review and Analysis [Internet]. Vol. 180, American Journal of Epidemiology. 2014. p. 865–75. Available from: http://dx.doi.org/10.1093/aje/kwu209

8. Fit probability distribution object to data - MATLAB fitdist [Internet]. [cited 2020 Mar 12]. Available from: https://www.mathworks.com/help/stats/fitdist.html

9. Li Q, Guan X, Wu P, Wang X, Zhou L, Tong Y, et al. Early Transmission Dynamics in Wuhan, China, of Novel Coronavirus-Infected Pneumonia. N Engl J Med [Internet]. 2020 Jan 29; Available from: http://dx.doi.org/10.1056/NEJMoa2001316

10. Kuk AYC, Ma S. The estimation of SARS incubation distribution from serial interval data using a convolution likelihood. Stat Med. 2005 Aug 30;24(16):2525–37.

11. Lipsitch M, Cohen T, Cooper B, Robins JM, Ma S, James L, et al. Transmission dynamics and control of severe acute respiratory syndrome. Science. 2003 Jun 20;300(5627):1966–70.

12. Cowling BJ, Park M, Fang VJ, Wu P, Leung GM, Wu JT. Preliminary epidemiological assessment of MERS-CoV outbreak in South Korea, May to June 2015 [Internet]. Vol. 20, Eurosurveillance. 2015. Available from:http://dx.doi.org/10.2807/1560-7917.es2015.20.25.21163

13. Park SH, Kim Y-S, Jung Y, Choi SY, Cho N-H, Jeong HW, et al. Outbreaks of Middle East Respiratory Syndrome in Two Hospitals Initiated by a Single Patient in Daejeon, South Korea. Infect Chemother. 2016 Jun;48(2):99–107.

14. Jung S-M, Akhmetzhanov AR, Hayashi K, Linton NM, Yang Y, Yuan B, et al. Real time estimation of the risk of death from novel coronavirus (2019-nCoV) infection: Inference using exported cases [Internet]. Available from: http://dx.doi.org/10.1101/2020.01.29.20019547

15. Tuite AR, Fisman DN. Reporting, Epidemic Growth, and Reproduction Numbers for the 2019 Novel Coronavirus (2019-nCoV) Epidemic [Internet]. Annals of Internal Medicine. 2020. Available from: http://dx.doi.org/10.7326/m20-0358

16. Wu JT, Leung K, Leung GM. Nowcasting and forecasting the potential domestic and international spread of the 2019-nCoV outbreak originating in Wuhan, China: a modelling study. Lancet [Internet]. 2020 Jan 31; Available from: http://dx.doi.org/10.1016/S0140-6736(20)30260-9

17. Zhao S, Gao D, Zhuang Z, Chong M, Cai Y, Ran J, et al. Estimating the serial interval of the novel coronavirus disease (COVID-19): A statistical analysis using the public data in Hong Kong from January 16 to February 15, 2020 [Internet]. Epidemiology. medRxiv; 2020. Available from: https://www.medrxiv.org/content/10.1101/2020.02.21.20026559v1.abstract

18. Nishiura H, Linton NM, Akhmetzhanov AR. Serial interval of novel coronavirus (2019-nCoV) infections [Internet]. Infectious Diseases (except HIV/AIDS). medRxiv; 2020. Available from: https://www.medrxiv.org/content/10.1101/2020.02.03.20019497v1

19. Zhanwei Du, Xiaoke Xu, Ye Wu, Lin Wang, Benjamin J. Cowling, Lauren Ancel Meyers. Serial Interval of COVID-19 among Publicly Reported Confirmed Cases. Emerging Infectious Disease journal [Internet]. 2020;26(6). Available from: https://wwwnc.cdc.gov/eid/article/26/6/20-0357_article

20. Kenah E, Lipsitch M, Robins JM. Generation interval contraction and epidemic data analysis. Math Biosci. 2008 May;213(1):71–9.

